# Racial disparities, environmental exposures, and SARS-CoV-2 infection rates: A racial map study in the USA

**DOI:** 10.1101/2023.04.17.23288622

**Authors:** Wenyan Xu, Bin Jiang, Chris Webster, William C. Sullivan, Yi Lu, Na Chen, Zhaowu Yu, Bin Chen

**Author notes:** The authors contributed equally to this work (Wenyan Xu, Bin Jiang). Corresponding authors: Bin Jiang Postal Address: 614 Knowles Building, Pokfulam Road, The University of Hong Kong, Hong Kong SAR Bin Chen Postal Address: Division of Landscape Architecture, Faculty of Architecture, The University of Hong Kong, Hong Kong SAR.

## Abstract

Since the onset of the COVID-19 pandemic, researchers mainly examined how socio-economic, demographic, and environmental factors are related to disparities in SARS-CoV-2 infection rates. However, we don’t know the extent to which racial disparities in environmental exposure are related to racial disparities in SARS-CoV-2 infection rates. To address this critical issue, we gathered black vs. white infection records from 1416 counties in the contiguous United States. For these counties, we used 30m-spatial resolution land cover data and racial mappings to quantify the racial disparity between black and white people’s two types of environmental exposure, including exposures to various types of landscape settings and urban development intensities. We found that racial disparities in SARS-CoV-2 infection rates and racial disparities in exposure to various types of landscapes and urban development intensities were significant and showed similar patterns. Specifically, less racial disparity in exposure to forests outside park, pasture/hay, and urban areas with low and medium development intensities were significantly associated with lower racial disparities in SARS-CoV-2 infection rates. Distance was also critical. The positive association between racial disparities in environmental exposures and racial disparity in SARS-CoV-2 infection rates was strongest within a comfortable walking distance (approximately 400m).

**Highlights:**

- Racial dot map and landcover map were used for population-weighted analysis.
- Racial disparity in environmental exposures and SARS-CoV-2 infection were linked.
- Forests outside park are the most beneficial landscape settings.
- Urban areas with low development intensity are the most beneficial urban areas.
- Landscape and urban exposures within the 400m buffer distances are most beneficial.

## 1 Introduction

### 1.1 Background

Racial disparity in health outcomes is a critical challenge in the United States and many other countries (Braveman et al., 2011). Many studies suggest that socio-economic and demographic disparities can lead to environmental disparities which then lead to human health disparities (Song et al., 2021; World Health Organization, 2010). A handful of studies have suggested that racial disparities in SARS-CoV-2 infection rates have a significant relationship with the racial disparities in environmental exposure to various landscape and urban settings; However, there is still much we do not know (Lu, Chen, et al., 2021). This study investigates the relationships among racial disparities in landscape and urban environment exposure and disparities in SARS-CoV-2 infection rates. This research may help governments prioritize resources to provide more equitable access to supportive natural and urban settings to reduce racial disparities in health outcomes for current COVID-19 pandemic and similar public health crisis in the future.

Generally, previous studies exploring the disparities between environmental exposures and SARS-CoV-2 infection rates has considered four types of relationships although those studies have many limitations in research design and measurements:

(1) The relationship between landscape exposure and SARS-CoV-2 infection rates;
(2) The relationship between racial disparity in landscape exposure and racial disparity in SARS-CoV-2 infection rates;
(3) The relationship between exposure to urban settings or various urban densities and SARS-CoV-2 infection; and
(4) The relationship between racial disparity in exposure to urban settings and urban densities and racial disparities in SARS-CoV-2 infection rates.

### 1.2 Landscape exposure and SARS-CoV-2 infection

Landscape exposure can be defined as passive or active contact with various types of green space. A growing number of studies have explored the relationship between landscape exposure and SARS-CoV-2 infection rates. Generally, these studies suggest that higher levels of landscape exposure are associated with a lower risk of SARS-CoV-2 infection. One study found that county-level SARS-CoV-2 infection rates decreased for every 1% increase in tree canopy, suggesting green spaces’ protective effect (Klompmaker et al., 2021). Greater access to green spaces was associated with lower transmission speed of COVID-19 (Alidadi & Sharifi, 2022), and total green space was negatively associated with SARS-CoV-2 infection rate (Jiang et al., 2022) and mortality rate (Yang et al., 2022). In contrast, higher density of public landscape settings in a high-density city was associated with higher rates of COVID-19 morbidity during the early stages of the COVID-19 pandemic, perhaps because of increased social contact with other people elicited by those settings (You et al., 2020).

### 1.3 Racial disparity in landscape exposure and racial disparity in SARS-CoV-2 infection

Although studies exploring racial disparity in landscape exposure and racial disparity in SARS-CoV-2 infection rates are scarce, more general studies exploring disparities in landscape exposure and disparities in public health outcomes suggest a positive relationship. Researchers have found positive causal and correlational associations between disparities in landscape exposure and disparities in health outcomes. In a natural experiment study, patients with more visual contact with green landscapes had a better recovery from a standard surgery (Ulrich, 1984). In an onsite experimental study, adding renovated green spaces to neighborhoods reduced incidences of gun violence, crimes, and sense of fear (Branas et al., 2018). A statewide study in the United States found that a lower disparity in greenness in residential environments was linked to lower cardiovascular mortality disparities between black and white men (Iyer et al., 2020). A nationwide study in England found that, in greener areas, lower disparities in all-cause mortality and mortality from circulatory diseases were observed among different income groups (Mitchell & Popham, 2008).

A handful of studies suggest that greater racial disparity in landscape exposure may lead to a greater disparity in SARS-CoV-2 infection, but these studies have research design and measurement limitations. For example, one recent study found that a higher ratio of green spaces was linked to lower racial disparity in SARS-CoV-2 infection rates for the 135 most urbanized counties in the United States (Lu, Chen, et al., 2021). However, this study did not investigate the relationship in less urbanized counties. It also did not control for the spatial distribution of racial populations and green spaces in the statistical analysis. Another study found that racial disparity in access to greenness was associated with a lower incidence of COVID-19 infection in the United States (Spotswood et al., 2021). However, the study examined only 17 states and used Normalized Difference Vegetation Index (NDVI) to measure greenness, which cannot fully represent the nationwide association and differentiate the contributions of distinct types of landscape settings. Lastly and most importantly, both studies did not consider the spatial distribution of different green spaces and the spatial distribution of different racial populations, which make the findings vulnerable to a possible ecological fallacy. An ecological fallacy is a logical error that occurs when the characteristics of a group are attributed to an individual; individual attributes cannot be deduced from statistical inferences of the group to which such individuals belong (Firebaugh, 2001; Woodruff et al., 2018).

### 1.4 Urban exposure and SARS-CoV-2 infection

Urban exposure can be defined as daily exposure to urban settings with low, moderate, or high development intensity. Many studies have shown that exposure to settings with different levels of urbanicity can produce different impacts on public health (e.g., McMichael, 2000; Son et al., 2020). Studies have found that exposure to more urbanized environments is generally more harmful for both mental and physical health due to lack of access to green spaces (Das et al., 2008; Jiang et al., 2022; Luo & Jiang, 2022; Venkatramana & Reddy, 2002). However, other studies have found that developed urban environments have higher levels of social security, collaboration, and vitality, which may collectively promote residents’ general health (Jauho & Helén, 2022; Mouratidis & Poortinga, 2020). Few studies have explored how exposure to urban settings is related to airborne infectious disease rates. Moreover, whether and to what extent the collinearity effect does exists between landscape and urban exposures should be investigated so we can fully understand their effects on SARS-CoV-2 infection rates (Jiang et al., 2022; Paköz & Işık, 2022).

The few published studies investigating the relationship between various indicators of urbanicity and risk of SARS-CoV-2 infection show significant positive correlations between urban density and the number of SARS-CoV-2 infection or mortality rates (e.g., Jamshidi et al., 2020; Lin et al., 2020; Zhang & Schwartz, 2020). Less dense urban areas may have a lower risk of SARS-CoV-2 infection because they provide open spaces that allow for safer social distances and reduced density of ambient particles that convey the virus (Jiang et al., 2022; Lin et al., 2020; Wang et al., 2021). However, further investigation beyond the general linear association is necessary to fully understand the relationship. For example, a county or census tract may contain numerous parcels with different levels of development intensity and land cover types. A simple linear association may fail to describe the complex relationship.

### 1.5 Racial disparity in urban exposure and racial disparity in SARS-CoV-2 infection

To our best knowledge, very few studies have investigated the relationship between racial disparity in urban exposure and racial disparity in SARS-CoV-2 infection. One study investigated this issue for the most urbanized counties in the US, and three measures of urban exposure were removed due to the collinearity problem (Lu, Chen, et al., 2021) . This study did not explore the relationship for areas with a much larger range of urban exposure. Another study exploring the relationship used NDVI to measure greenness, but NDVI can only roughly indicate the level of urban development intensity. More accurate measures are needed (Spotswood et al., 2021). Lastly and most importantly, both studies did not consider the spatial distribution of land parcels with different levels of development intensity and the spatial distribution of different racial populations, which make the findings may have a high risk of ecological fallacy.

### 1.6 Critical knowledge gaps

In our understanding of the relationship between racial disparity in landscape and urban exposure and racial disparity in SARS-CoV-2 infection rates, five gaps remain.

First, previous studies do not consider the spatial distribution of racial populations. This makes it difficult to differentiate their exposure to nearby landscape and urban settings and leaves the findings vulnerable to ecological fallacy.

Second, previous cross-sectional studies on racial disparity issues do not adopt a within-county comparison between races. A within county comparison between racial populations may reduce the probability of ecological fallacy by mitigating bias caused by a wide variety of distinct conditions among different zones (Lu, Chen, et al., 2021).

Third, previous studies use NDVI or total greenness as measures of landscape exposure and are unable to differentiate the effects of different types of green spaces.

Fourth, few studies have comprehensively measured urban exposure, leading to a gap in our understanding of the association between urban exposure (exposure to different levels of development intensity) and racial disparity in SARS-CoV-2 infection rates.

Last, previous studies have only examined highly urbanized counties or a few regions. These findings may fail to describe the relationship between racial disparity in landscape and urban exposure and racial disparity in SARS-CoV-2 infection rates on a nationwide scale or in countries with various levels of urban density.

### 1.7 Research questions

To address these knowledge gaps, we conducted a one-year nationwide study by adopting high-resolution land cover and racial population maps. We asked the following five questions:

(1) Whether and to what extent did black and white people have significant differences in SARS-CoV-2 infection rates?
(2) Whether and to what extent did black and white people have significant differences in landscape and urban exposures?
(3) Whether and to what extent were landscape and urban exposures significantly associated with infection rates for black and white people?
(4) Whether and to what extent was the racial disparity in landscape and urban exposures significantly associated with racial disparity in SARS-CoV-2 infection rates?
(5) Whether and how did the association between racial disparity in landscape and urban exposures and SARS-CoV-2 infection rates change as the buffer distance changed?

## 2 Method

We adopted a within-county research design with 1416 counties in the contiguous United States (US). The COVID-19 case data were collected from January 1, 2020, to December 31, 2020. In this study, the racial disparities were measured as the difference in environmental exposures and infection rates between black and white populations in the same county. The methods included defining study areas; calculating SARS-CoV-2 infection rates, landscape and urban exposures, and socio-economic and demographic characteristics; and finally conducing statistical analysis.

### 2.1 Study areas

We used counties, the fundamental administrative unit in the United States, as the basic unit of analysis. Using publicly available data from the US Centers for Disease Control and Prevention (Centers for Disease Control and Prevention, 2021), we collected infection case data and associated race and ethnicity information from a total of 3,108 counties. We excluded 1,691 counties that did not have race records for black and/or white groups (452 counties had no records for white groups, 1691 counties had no records for black groups, and 452 counties had no records for both black and white groups). We also excluded La Salle County in Texas because of a data collection error. 1416 counties were included in the study.

1223 of the 1416 selected counties (86.4%) had at least 2% black population (1633 of the 3142 counties in the US have more than 2% black population). Most of the unselected counties (those that did not have race records for black and/or white groups) contained too low of a ratio of black people. In addition, all selected 1416 counties had more than 6% white people.

To further evaluate whether the chosen 1416 counties are representative of the contiguous United States, we applied a random selection strategy based on geographic spatial distribution to select sample counties for sensitivity analysis. The country was divided into nine divisions by the United States Census Bureau. The average rate of sample availability (1416/3142) was set as the threshold to randomly select samples in each of the nine divisions. If the available counties in the division were less than the threshold, all the available counties were selected. Otherwise, we used random sampling methods with the threshold value. We applied this random selection strategy five times to generate five sample groups. Then, we analyzed the data from these five groups of counties to answer the research questions and found high agreement in our results. Detailed findings can be found in Appendix B. Therefore, we argue that the sample pool of 1416 counties is representative enough for us to analyze the relationship between racial disparity in landscape and urban exposure and racial disparity in SARS-CoV-2 infection rate in the contiguous United States.

### 2.2 SARS-CoV-2 infection rates

To calculate the SARS-CoV-2 infection rate for black and white people, we retrieved the total white and black population counts for each county, retrieved from 2019 census data (United States Census Bureau, 2020). The racial disparity in SARS-CoV-2 infection rates was calculated as the difference between the infection rate in black individuals and the infection rate in white individuals for the same county.

### 2.3 Landscape and urban exposures: Land cover & racial population mapping

#### 2.3.1 Landscape settings and urban areas with different levels of development intensity

The National Land Cover Database (NLCD) is an operational land cover monitoring program providing updated land cover and related information for the US. Using the NLCD 2019 dataset (https://www.mrlc.gov), we extracted two major types of land cover: landscape settings (green spaces) and urban areas with different levels of development intensity. The following land-cover types were classified as landscape settings: a) developed open space; b) deciduous forest, evergreen forest, and mixed forest (combined into one forest category); c) grassland and herbaceous; and d) pasture and hay. Urban areas were classified according to low, medium, or high development level based on the proportion of constructed material and vegetation (Yang et al., 2018) . Low development intensity is typified by mostly vegetation (20–49% impervious surfaces), medium development intensity by single-family homes (50– 79% impervious surfaces), and high development intensity by frequently traversed areas (80– 100% impervious surfaces).

#### 2.3.2 Mapping racial populations in the United States

We used racial data collected and shared by SocScape (Social Landscape) to map the black and white populations in the United States at a high resolution (30m grids) (Figure 1) SocScape is a research project which aims to provide mapping resources to visualize and analyze residential segregation and racial diversity in the conterminous United States and United States metropolitan areas (http://www.socscape.edu.pl/). It shows 30m grids of seven race/ethnicity sub-populations: non-Hispanic whites, non-Hispanic blacks, non-Hispanic Asians, non-Hispanic American Indians, non-Hispanic Native Hawaiians and Other Pacific Islanders, non-Hispanic other races, and Hispanics. The racial dot map of non-Hispanic whites and non-Hispanic blacks was used in this study (Figure 1). The 30m grids have been developed using dasymetric modeling, which has a high accuracy of gridded population data over commonly used Census tract/blocks-aggregated data (Dmowska & Stepinski, 2019; Dmowska et al., 2017).

**Figure 1.**
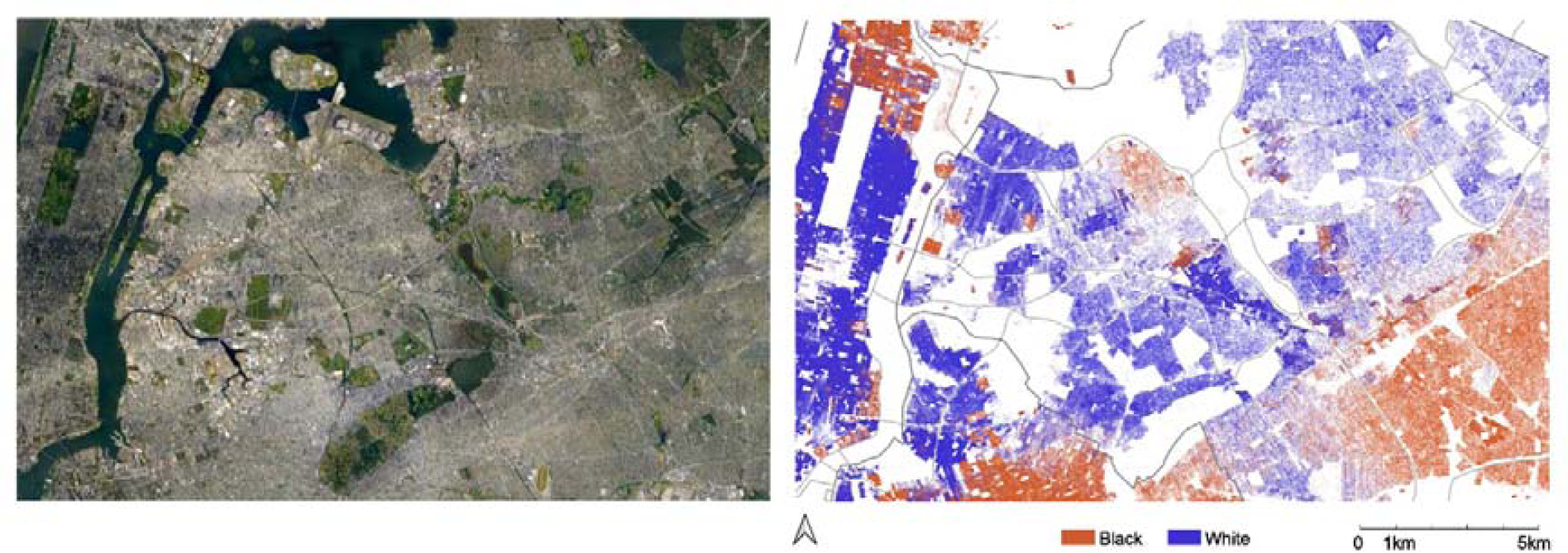
Left: The Google Earth image of an area in the New York City, USA; Right: The black-white racial dot map of the same area.

#### 2.3.3 Exposure to landscape settings

The four types of landscape settings with predominant natural elements were assessed at 30m resolution. We divided developed open space and forest by park boundaries into inside and outside park factors (Ersi, 2021). We calculated black-white population-weighted green space exposure for varying buffer distances in each county to identify the impact of different landscape settings on racial disparity in infection rates. The population-weighted exposure assessment incorporated the spatial distribution of population footprint into landscape exposure estimates by giving proportionally greater weight to landscape settings near areas with higher densities of human residents.

Race population-weighted green space exposure was calculated with the Google Earth Engine (GEE) using NLCD 2019. The 30m resolution NLCD, 2019 Landsat imagery matched well with the 30m spatial resolution of the racial map in GEE. This enabled the population-weighted landscape exposure within various buffer sizes in each county. The race population-weighted landscape exposure for different buffer sizes in each county is defined in Equation (1) (Chen, Song, Jiang, et al., 2018; Chen, Song, Kwan, et al., 2018; Chen et al., 2022),

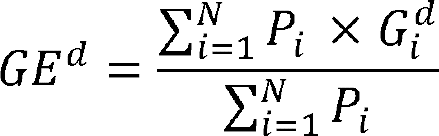

where *P_i_* is the black or white population of the *i*th grid, G^d^ is the green space of the *i*th grid at a buffer size of *b* meters, *N* is the total number of grids for a given county, and *GE* is the estimated landscape exposure level for the given county. We calculated landscape exposure levels by varying the buffer size from 100m to 5km (the maximum appropriate walking distance in the United States, based on Yang & Diez-Roux, 2012) to examine the association between different proximities to landscape settings and the infection disparity between black and white people. The relative exposure difference between black and white people of these areas was calculated as explanatory variables in the generalized linear mixed model to examine their contribution to mitigating racial disparity of infection rates.

#### 2.3.4 Exposure to areas with different levels of development intensity

The dominant land-cover types in the areas with different levels of development were also considered. We calculated the exposure ratio using the same methods we used for landscape settings. Urban areas with three levels of development intensity from NLCD 2019 were categorized as urban areas with low, medium, and high development intensity (Appendix Table S1(a)). The exposure difference between black and white people in these areas was also calculated using buffer sizes from 100m to 5km.

### 2.4 Socio-economic and demographic characteristics as covariates

Previous studies have shown that socioeconomic and demographic characteristics are important predictors of SARS-CoV-2 infection risk and racial disparity in infection rates (Abedi et al., 2020; Figueroa et al., 2020). Thus, we adjusted for the potential confounding factors of black or white people’s socioeconomic and demographic characteristics. The descriptive statistics for each variable are presented in Table 1 and Appendix Table S1 (b).

**Table 1.**
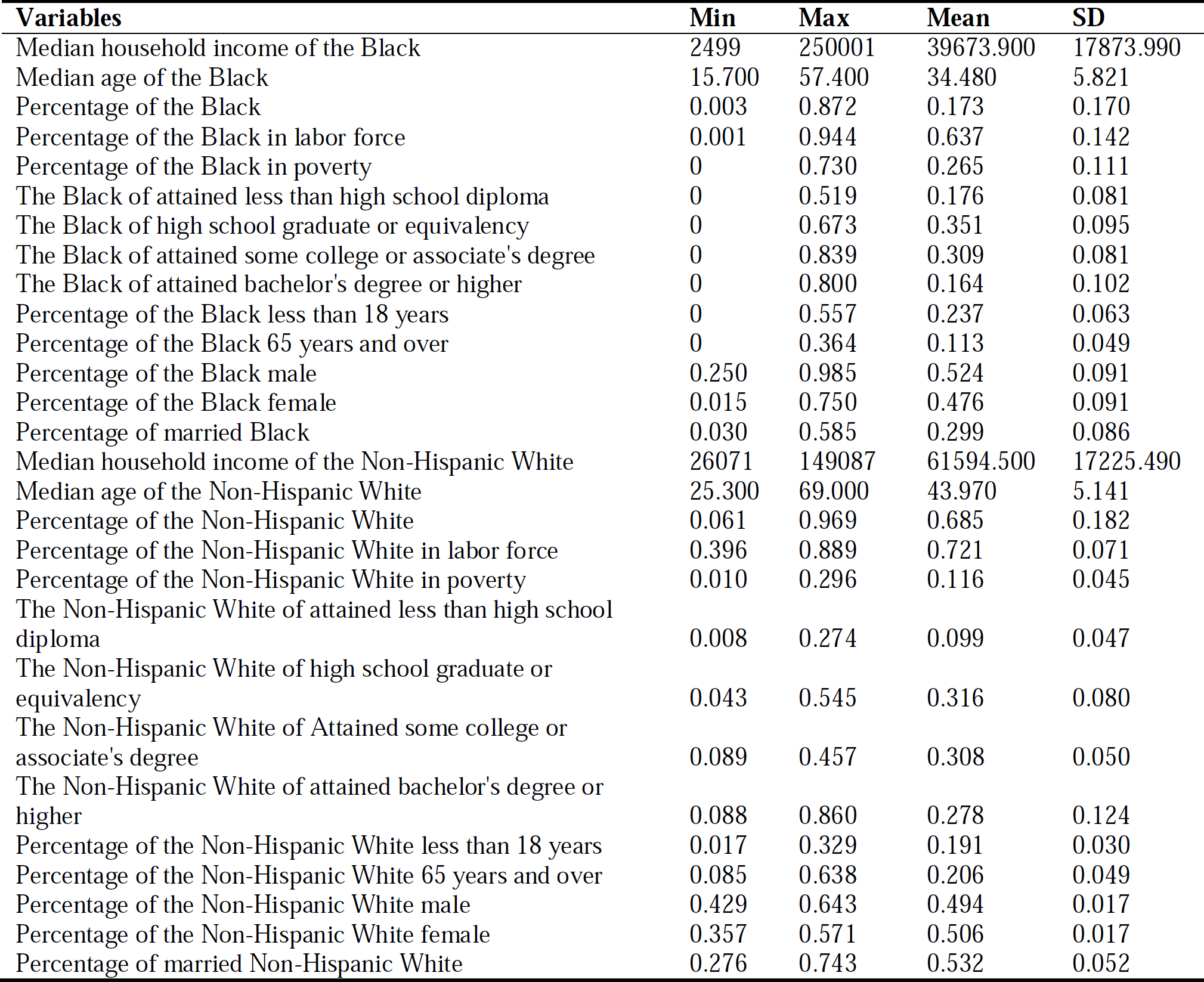
Descriptive statistics of socio-demographic characteristics of 1320 counties (96 counties without the information)

### 2.5 Statistical analysis

The main statistical analysis framework is presented in Figure 2. It includes five components. Each component aims to answer one research question proposed in Section 1.7.

**Figure 2.**
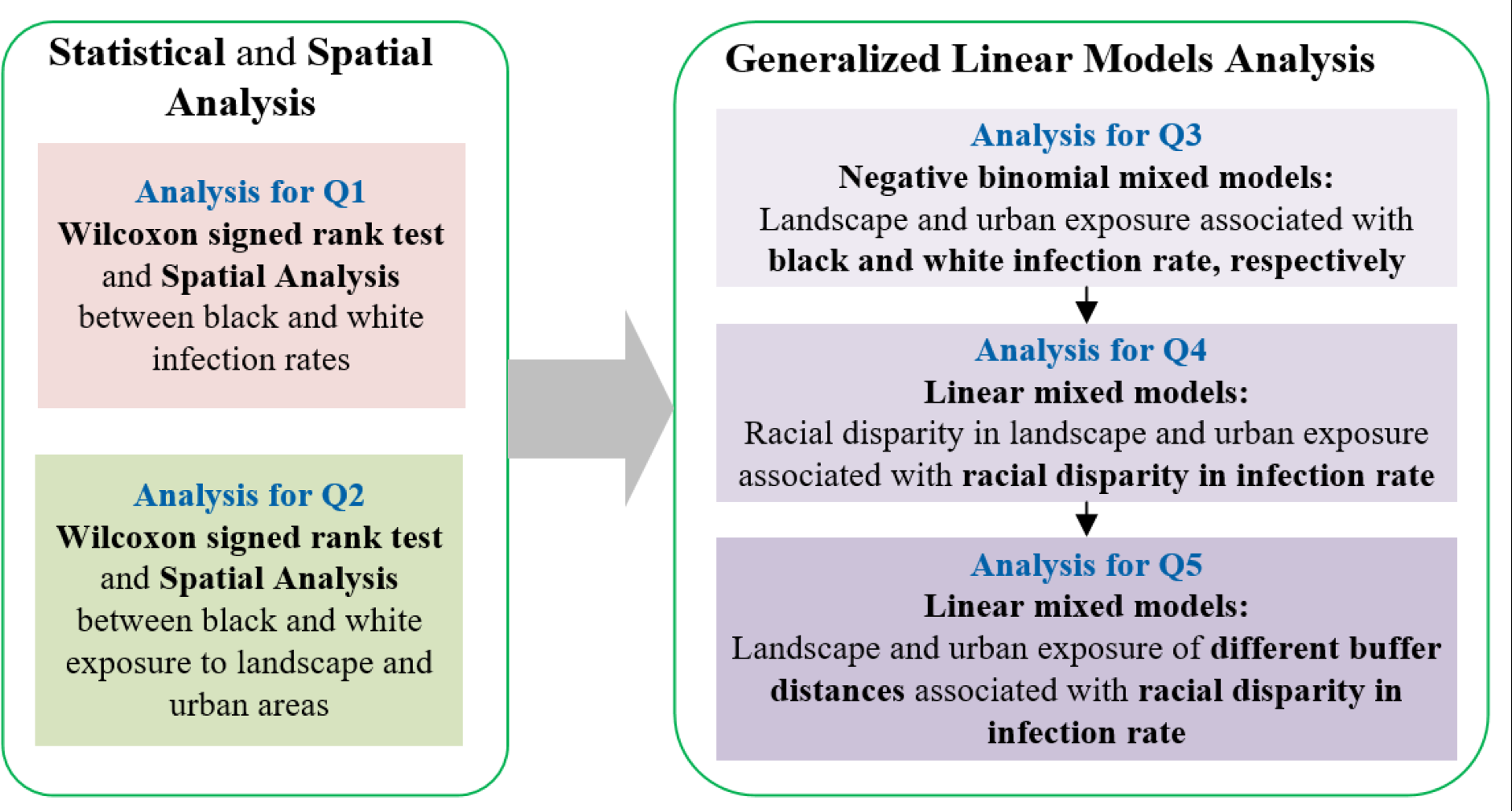
The framework of statistical analysis includes five components. Each component aimed to answer one of five proposed research questions.

First, we used the Wilcoxon signed rank test to examine whether there was a significant difference in SARS-CoV-2 infection rates between black and white people in the same county. We examined whether there was a significant difference between black and white people’s exposure to landscape settings and urban areas with different development intensities in the same county. Second, we applied negative binomial regression models to investigate associations between landscape and urban exposures and people’s SARS-CoV-2 infection rates. In the models, black infection rates and white infection rates were dependent variables, and socio-economic and demographic characteristics were covariates.

We also included a random intercept for states to account for the non-independence of data from the same state. Third, we adopted generalized linear mixed models to explore the extent to which the disparity of population landscape and urban exposure is related to racial disparity in SARS-CoV-2 infection rates, after accounting for covariates. In the models, the relative differences between black and white infection rates were the dependent variables, and exposures to various landscape settings and urban areas between black and white people were the independent variables, while difference of socioeconomic and demographic characteristics between the two groups were included as covariates. We also included a random intercept by state to account for a potential correlation in counties within the same state.

A buffer distance of 400m was used to measure population-weighted landscape and urban exposure in all above models, since this buffer distance can be regarded as a convenient distance for most walking trips for people in the United States (Yang & Diez-Roux, 2012), and a common walking distance between residential locations (300m to 500m) and landscape settings, recommended by the World Health Organization (2016).

To address the issue of spatial autocorrelation in these models, we built additional simultaneous autoregressive models (SAR) to validate the results of these generalized linear models. In the first, we confirmed the presence of spatial autocorrelation in the models and then used a SAR model to adjust for the presence of spatial autocorrelation. We used the queen criteria to build the neighbors matrix and model selection with Akaike information criterion (AIC) to compare three potential structures for where the spatial autoregressive process is believed to occur. These potential structures include: a spatial error model, where spatial dependence is assumed to occur in the error term; a spatial lag model, where spatial dependence is assumed to occur in the response variable; and a mixed or Durbin model, where spatial dependence is assumed to influence both the response and explanatory variables.

Finally, we used a series of generalized linear mixed models to explore the relationship between the difference of black and white infection rates and varying buffer distances to landscape settings and urban areas with different levels of development intensity. Apart from 100m, the buffer distances varied from 200m to 1800m by intervals of 200m. For buffer distances between 2km and 5km, we used intervals of 500m. We performed negative binomial mixed models and generalized linear mixed analyses using R v4.1.1 with built-in ‘glmer.nb’, and ‘lmer’ function, respectively (R Core Team, 2020). The inclusive variables for all models were refined with the variance inflation factor criterion (VIF) ≥ 4, to remove multicollinearity from the regression (O’brien, 2007). The model coefficient estimates, standard errors, degree of freedom, t-values, and p-values for coefficient estimates were reported.

## 3 Results

### 3.1 Whether and to what extent did a racial disparity in infection rates exist?

As of Dec. 31, 2020, there were a total of 4,370,477 cases among black and white people in the 1,416 sample counties of the United States (Figure 3a). The county erage infection rate for white individuals was 2,579 persons per 100,000, whereas the infection rate for black individuals was 3,171 per 100,000 (Figure 3b). The average black-white difference in infection rate was 592 persons per 100,000 population. White individuals had a higher infection rate than black individuals in only 483 out of the 1416 counties. As expected, a Wilcoxon signed rank test revealed that the difference in infection rates between black and white people was significant, *p* < 0.001.

**Figure 3.**
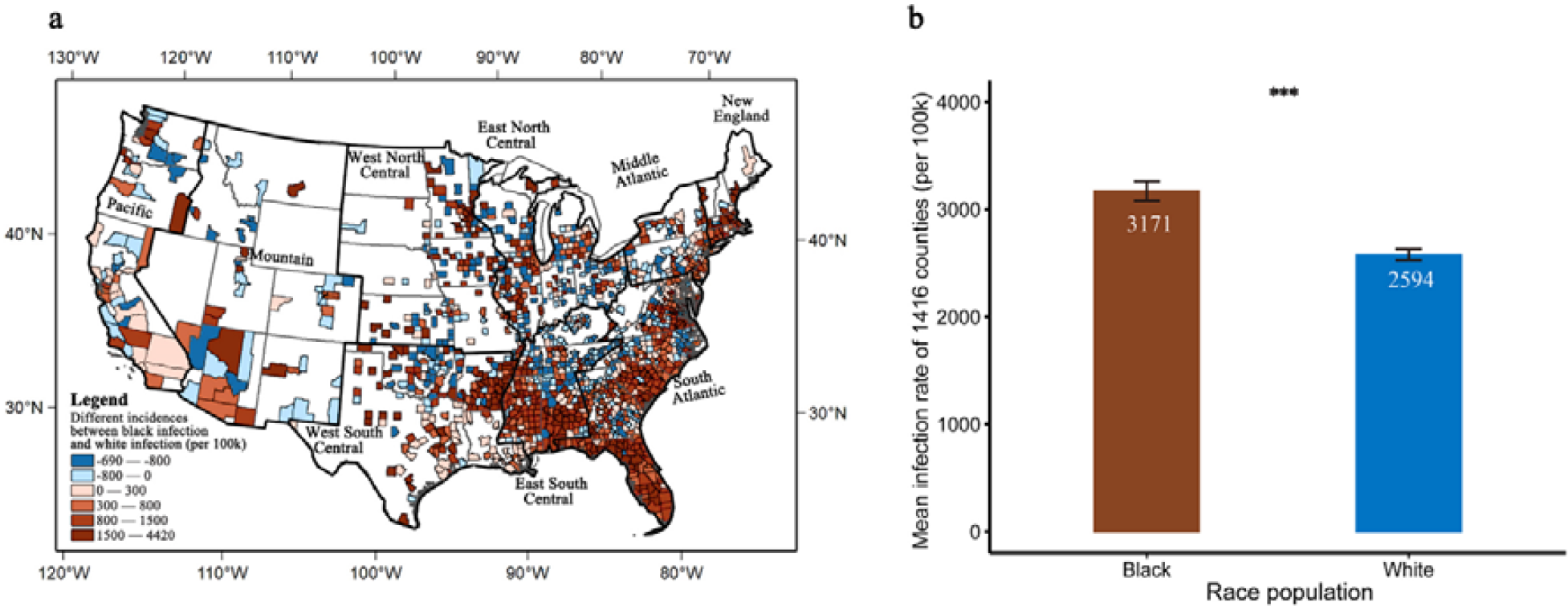
Difference in SARS-CoV-2 infection rates between black and white people. a. Difference in infection rates between black and white people within counties (per 100k population). Blue color indicates that white people have a higher infection rate than black people, and red color indicates that black people have a higher infection rate than white people. b. The mean SARS-CoV-2 infection rates between black and white population groups. The significant difference is identified at *p* < 0.001 using Wilcoxon signed rank test. The error bar represents standard error of the mean.

### 3.2 Whether and to what extent did racial disparities in landscape and urban exposures exist?

The Wilcoxon two-sample test found that, comparing with white people, black people had significantly lower exposure to multiple types of landscape settings, including forests, grassland/herbaceous, and pastures/hay. Additionally, black people had significantly higher exposure to urban areas with three different levels of development intensity (Figure 4 & Figure 5). The findings confirmed that there were significant and large racial disparities in landscape and urban exposure.

**Figure 4.**
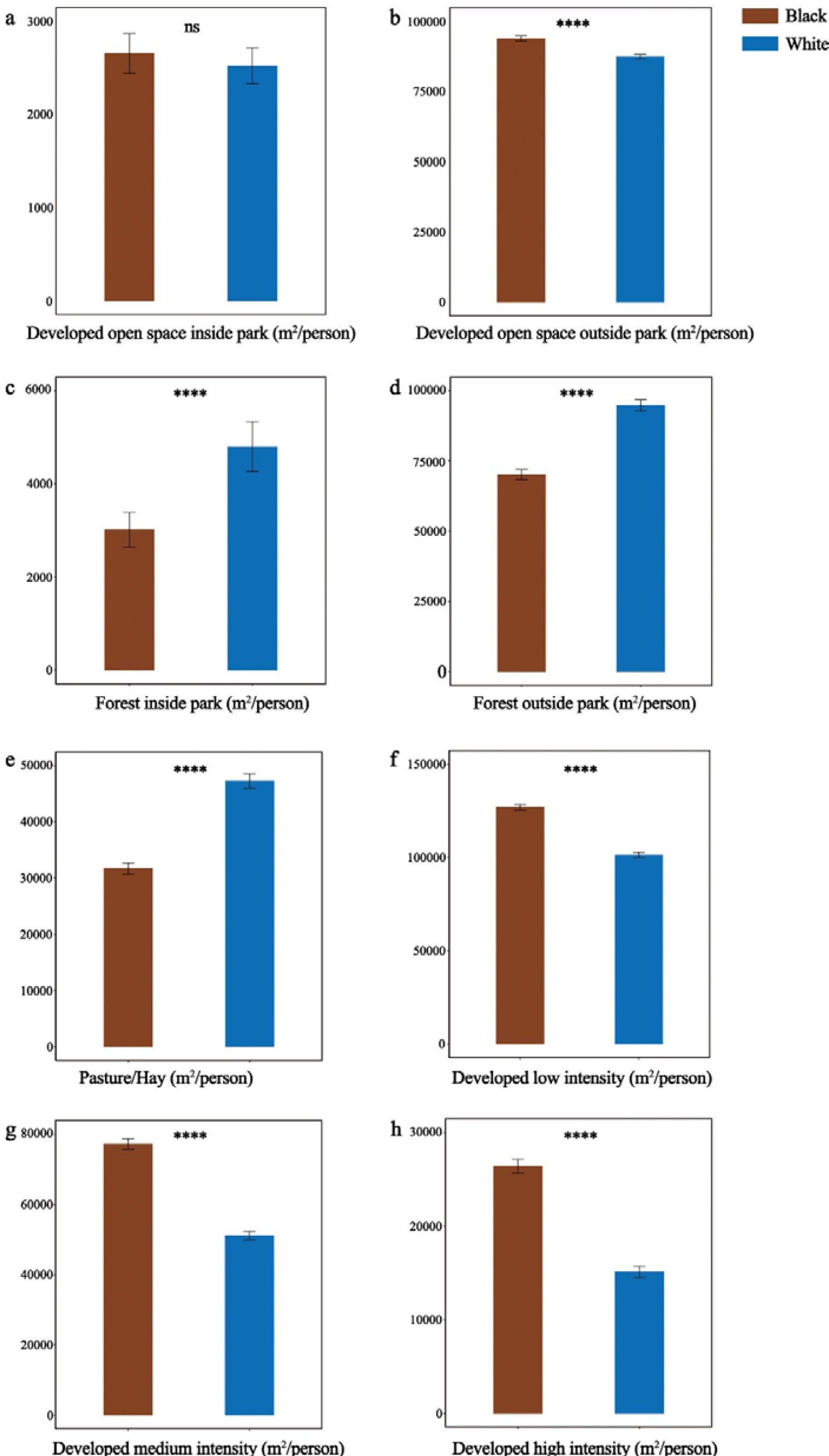
Comparisons of landscape and urban exposure between black and white people (within the 400 m buffer distance). A Wilcoxon two-sample test was used to compare eight types of landscape and urban exposure. The histogram shows mean values of exposure for black and white people in 1416 counties. The error bar represents standard error of the mean. ‘****’ *p* < 0.0001, ‘ns’ *p* ≥ 0.1.

**Figure 5.**
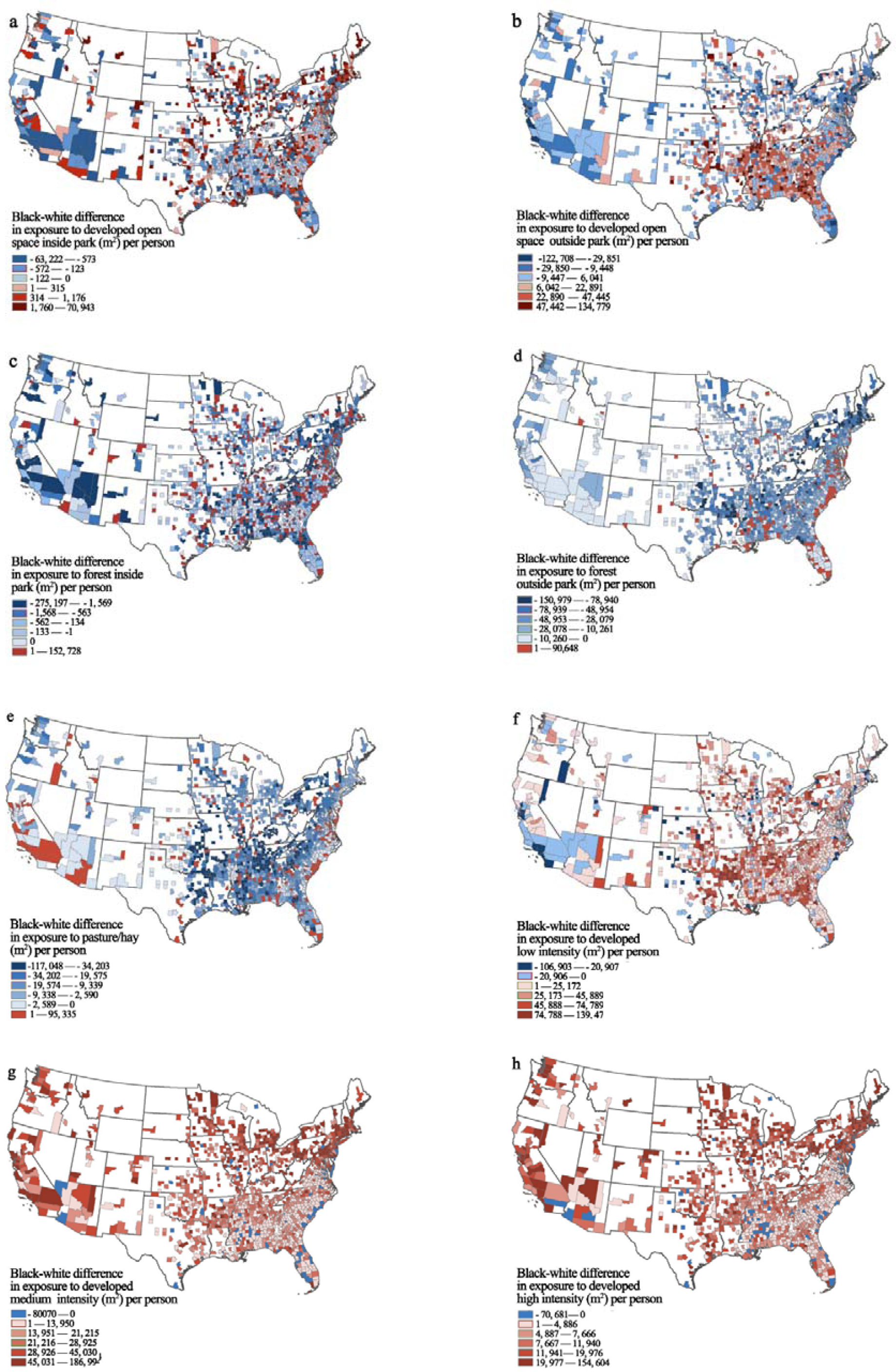
Within-county racial difference in landscape and urban exposure for each of 1416 counties (within the 400m buffer distance).

### 3.3 Whether and to what extent were landscape and urban exposures significantly associated with SARS-CoV-2 infection rates?

As shown in Figure 6, we identified several significant associations between landscape and urban exposure and SARS-CoV-2 infection rates for each race. For black people, both forest inside park and urban areas with medium development intensity yielded a significant and negative association with infection rates (*p* < 0.05). For white people, forest outside park yielded a significant and negative association with infection rates (*p* < 0.05). Urban areas with high development intensity yielded a significant and negative association with infection rates (*p* < 0.05) (see detailed results in Supplementary Table S2).

**Figure 6.**
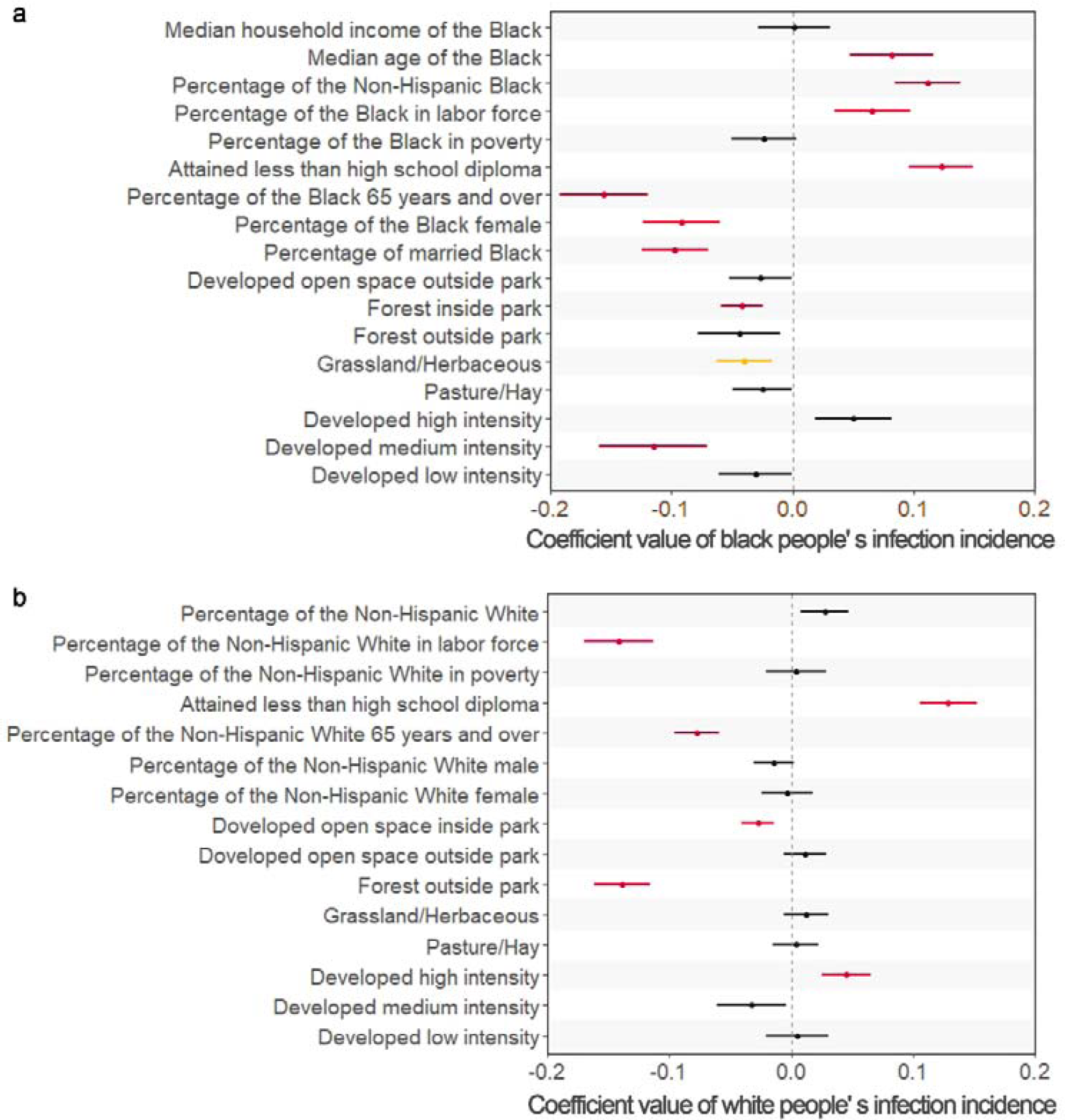
Coefficient values represent effect sizes from a negative binomial mixed effects model for the relationship between rates of COVID-19 black (a) or white (b) cases per 100,000 people, landscape exposure, urban exposure, and socioeconomic and demographic factors. The landscape and urban exposures were measured within the 400m buffer for each race. Coefficient values (dots and bars) represent 95% CIs. Significant variables are shown in red (*p* < 0.05), and marginally significant in yellow (*p* < 0.1) to present associations between black or white exposure to green spaces and areas with different levels of development intensity, and black or white infection rate.

**Table 2.** Model results from the generalized linear mixed effects model.

|  | VIF | Estimate | Std.<br>Error | df | t<br>value | Pr( > \|t\| ) |
| --- | --- | --- | --- | --- | --- | --- |
| (Intercept) |  | 0.264 | 0.06 | 38.15 | 4.18 | <0.001*** |
| Diff. in median household income | 1.49 | -0.046 | 0.02 | 1298 | -2.19 | 0.029* |
| Diff. in median age | 2.68 | 0.085 | 0.03 | 1311 | 2.70 | 0.007** |
| Diff. in percentage of population | 1.29 | 0.194 | 0.02 | 1319 | 8.32 | <0.001*** |
| Diff. in percentage of population in labor force | 2.28 | 0.087 | 0.03 | 1295 | 2.60 | 0.009** |
| Diff. in percentage of the population in poverty | 1.47 | -0.007 | 0.02 | 1299 | -0.31 | 0.76 |
| Diff. in attained less than high school diploma | 1.34 | 0.100 | 0.02 | 1303 | 4.16 | <0.001*** |
| Diff. in percentage of the population 65 years and over | 2.7 | -0.126 | 0.03 | 1310 | -3.80 | <0.001*** |
| Diff. in percentage of female | 2.36 | 0.017 | 0.04 | 1311 | 0.48 | 0.634 |
| Diff. in percentage of married population | 1.61 | -0.041 | 0.02 | 1312 | -1.68 | 0.093 |
| Diff. in developed open space inside park | 1.36 | -0.007 | 0.02 | 1281 | -0.35 | 0.728 |
| Diff. in developed open space outside park | 1.67 | -0.034 | 0.03 | 1288 | -1.33 | 0.183 |
| Diff. in Forest inside park | 1.35 | 0.005 | 0.03 | 1284 | 0.16 | 0.87 |
| Diff. in Forest outside park | 1.92 | -0.088 | 0.03 | 1317 | -3.37 | <0.001*** |
| Diff. in Grassland/Herbaceous | 1.15 | -0.019 | 0.02 | 1319 | -0.82 | 0.412 |
| Diff. in Pasture/Hay | 1.62 | -0.053 | 0.02 | 1311 | -2.21 | 0.027* |
| Diff. in high development intensity | 1.64 | 0.034 | 0.03 | 1312 | 1.30 | 0.195 |
| Diff. in medium development intensity | 1.93 | -0.073 | 0.03 | 1315 | -2.49 | 0.013* |
| Diff. in low development intensity | 1.94 | -0.073 | 0.03 | 1305 | -2.74 | 0.006** |

Note: Diff. means difference between black and white population. Both landscape exposure and urban exposure were measured within the 400 m buffer distance for each race.

### 3.4 Whether and to what extent were racial disparities in landscape and urban exposures significantly associated with racial disparity in infection rates?

The generalized linear mixed effects model revealed a significantly negative correlation between exposure to forest outside park and pasture and racial disparity in COVID-19 cases. As shown in Table 2 & Figure 7, we identified a strong association between greenspace exposure and racial disparity of SARS-CoV-2 infection rates. A unit difference of forests outside parks between black and white populations had a significant association with a decrease of 8.83% in the racial disparity in infection rates (*p* < 0.001). We found that the significant association of pasture/hay was associated with a decrease of 5.34% in the racial disparity in infection rates (*p* < 0.05). The difference in exposure to development intensity exerted a mediating effect on SARS-CoV-2 infection rates decided by the levels of intensity. Overall, population exposure to developed urban areas with low (β = -0.073, *p* < 0.01) and medium intensities (β = -0.073, *p* < 0.05) was significantly associated with low racial disparity of infection rates (Figure 7).

**Figure 7.**
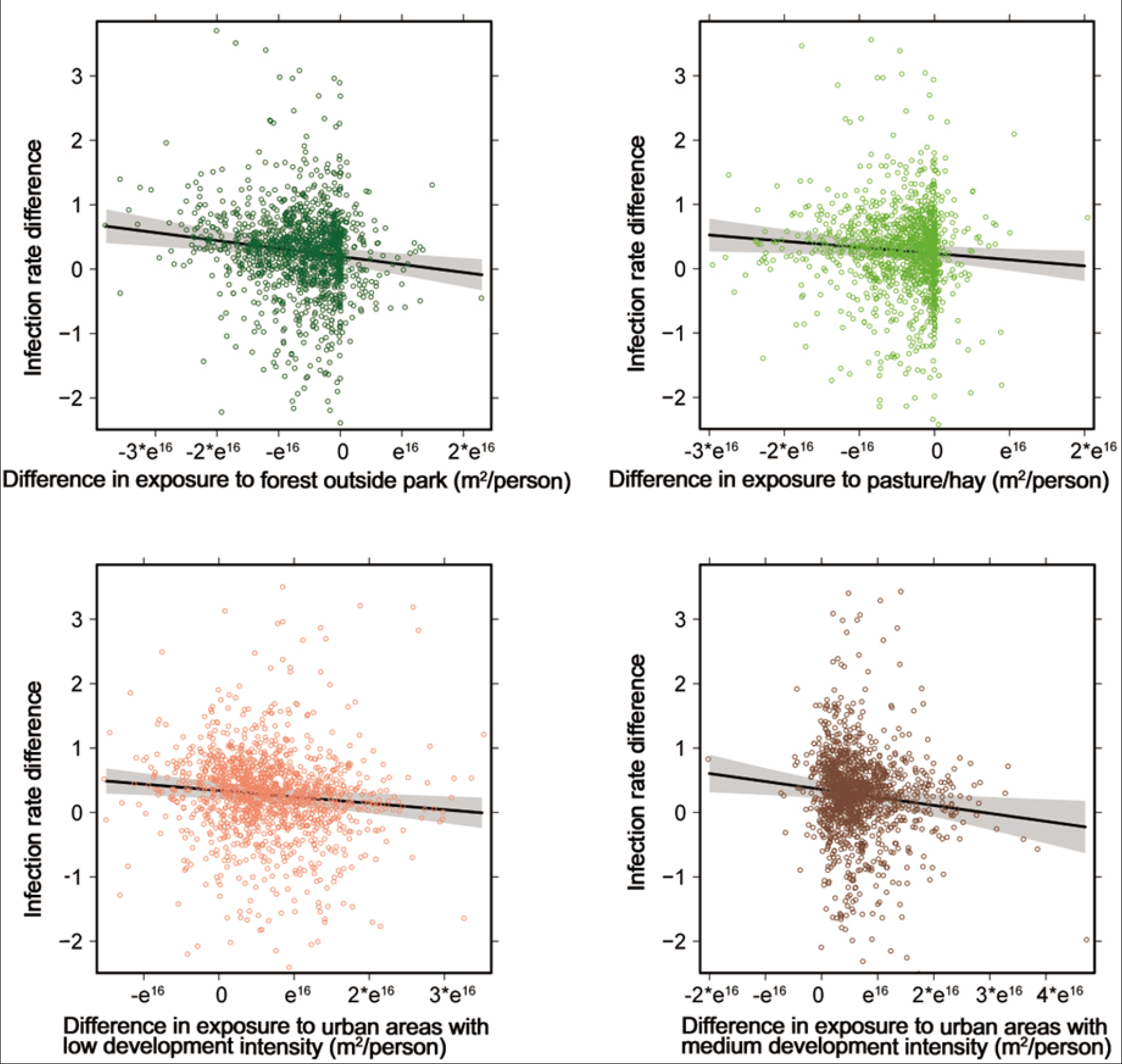
Green spaces and developed urban areas with developed intensities that have significant negative correlation with racial disparity of infection rate. The relative difference of infection rate between black and white population was calculated as ln (black cases/100k) - ln (white cases/ 100k).

### 3.5 Whether and how did the association between racial disparity in landscape and urban exposures and SARS-CoV-2 infection rates change as the buffer distance changed?

The results of sensitivity analysis of the four significant exposures found in 3.4 are shown in Figure 8 and Table 3. For the racial disparity in exposure to forest outside park, the buffer distance of best efficacy is 400m. The efficacy remains significant up until 1600m. For the racial disparity in exposure to pasture/hay, the buffer distance of best efficacy is 200m. The efficacy remains significant up until 400m. For the racial disparity in exposure to developed low intensity area, the buffer distance of best efficacy is 100m. The efficacy remains significant up until 600m. For the racial disparity in exposure to urban areas with moderate development intensity, the buffer distance of best efficacy is 600m, and the efficacy remains significant up until 1200m.

**Figure 8.**
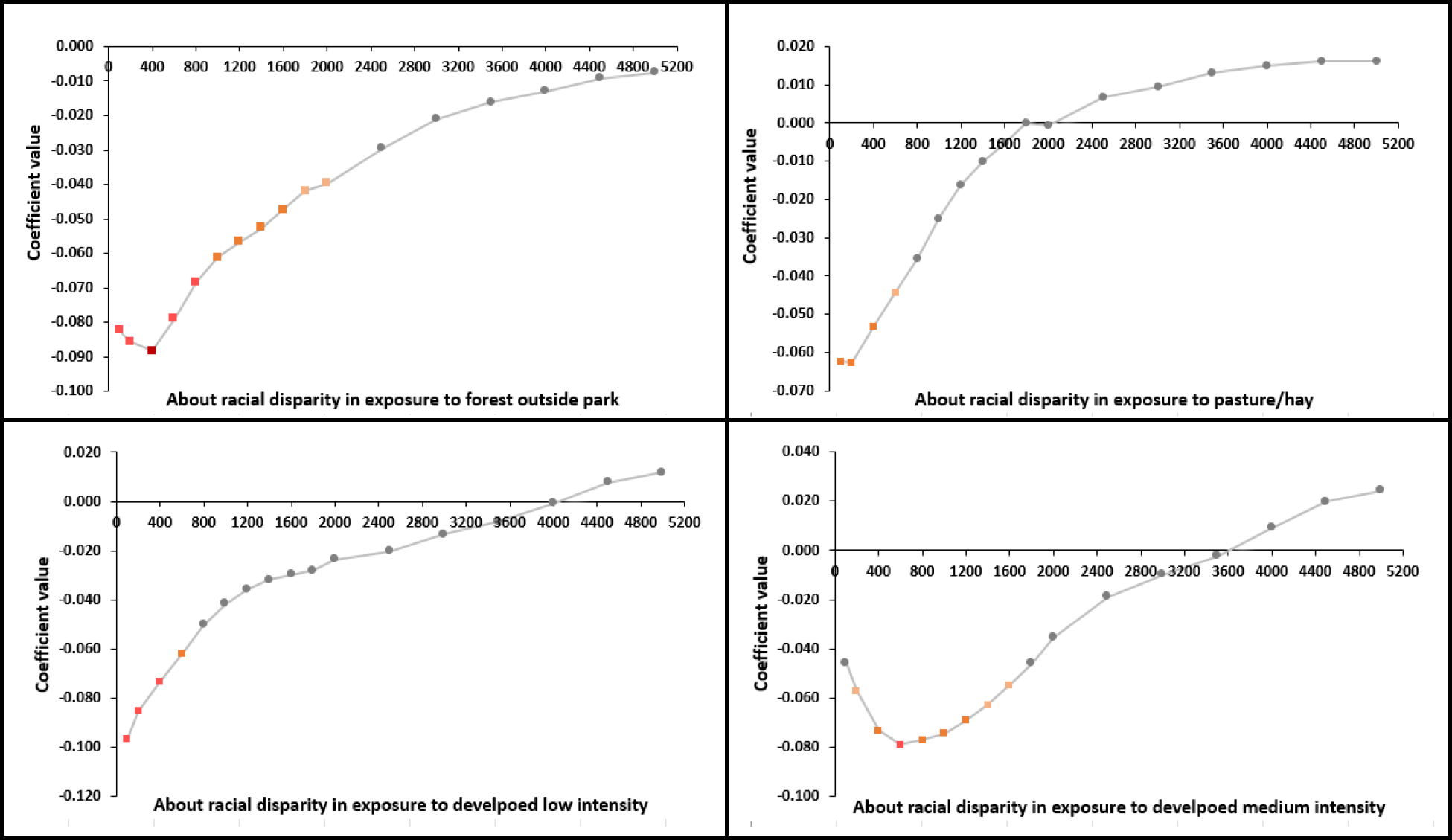
Coefficient values describing the associations between racial disparity in landscape and urban exposure within multiple buffer distances (100m-5km) and racial disparity in SARS-CoV-2 infection rates. Coefficient values are represented as dots with different colors. The gray color indicates nonsignificant associations, *p* ≥ 0.1. Strong to light red colors indicate significant associations at different levels, *p* < 0.001, *p* < 0.01, *p* < 0.05, and *p* < 0.1 (see detailed numbers in Table 3).

**Table 3.** Coefficient values describing the associations between racial disparity in landscape and urban exposure within multiple buffer distances (100m-5km) and racial disparity in SARS-CoV-2 infection rates. Significant levels are at ^ns^p ≥ 0.1, ^†^p < 0.1, *p < 0.05, **p < 0.01, and ***p < 0.001.

| Buffer distance<br>(meter) | Landscape exposure |  | Urban exposure |  |
| --- | --- | --- | --- | --- |
|  | Forest outside<br>park | Pasture/hay | Medium development<br>intensity | Low development<br>intensity |
| 100 | -0.082** | -0.062* | -0.046 | -0.097** |
| 200 | -0.086** | -0.063* | -0.057† | -0.085** |
| 400 | -0.088*** | -0.053* | -0.073* | -0.073** |
| 600 | -0.079** | -0.044† | -0.079** | -0.062* |
| 800 | -0.068** | -0.035 | -0.077* | -0.050† |
| 1000 | -0.061* | -0.025 | -0.074* | -0.042 |
| 1200 | -0.057* | -0.016 | -0.069* | -0.036 |
| 1400 | -0.053* | -0.010 | -0.063† | -0.032 |
| 1600 | -0.047* | -0.005 | -0.055† | -0.030 |
| 1800 | -0.042† | 0.000 | -0.046 | -0.028 |
| 2000 | -0.040† | -0.001 | -0.036 | -0.024 |
| 2500 | -0.030 | 0.007 | -0.019 | -0.020 |
| 3000 | -0.021 | 0.010 | -0.010 | -0.013 |
| 3500 | -0.016 | 0.013 | -0.002 | -0.008 |
| 4000 | -0.013 | 0.015 | 0.009 | -0.001 |
| 4500 | -0.009 | 0.016 | 0.020 | 0.008 |
| 5000 | -0.008 | 0.016 | 0.024 | 0.012 |

In total, all measures achieve a significant efficacy when the buffer distance is within 400m, which proves the validity of choosing 400m as the buffer distance for analysis in previous sections. Moreover, the beneficial associations gradually diminish as buffer distance increases. All become nonsignificant when the buffer distance is greater than 1600m.

## 4 Discussion

We present three major findings: (1) Racial disparity is profound: racial disparity in SARS-CoV-2 infection rates and racial disparity in landscape and urban exposure were significant and showed similar patterns. (2) Landscape and urban exposures impact racial disparity in SARS-CoV-2 infection rates. Less racial disparity in exposure to forests outside park and pasture/hay and urban areas with low and medium development intensities were significantly associated with lower racial disparities in SARS-CoV-2 infection rates. (3) Distance is critical: landscape and urban exposure within walking distance (around 400m) was significantly associated with racial disparity in SARS-CoV-2 infection rates.

The following discussion provides interpretations and implications of these major findings. We report limitations and recommend opportunities for future research.

### 4.1 Why was less racial disparity in landscape exposure associated with less racial disparity in infection rates?

**We describe four mechanisms that may explain the association.** First, less racial disparity in landscape exposure may lead to less disparity in air pollution exposure, which may contribute to lower racial disparity in infection rates. Recent studies have shown that air pollution is positively associated with higher COVID-19 mortality rates (Wu et al., 2020) and COVID-19 infection rates (Zhu et al., 2020) because COVID-19 is an airborne disease (Klompas et al., 2020). Studies found that a higher density of PM2.5, PM10, NO2, or O3 in the air was linked to a higher SARS-CoV-2 infection rate (Zhu et al., 2020). Less racial disparity in landscape exposure may reduce racial disparity in air pollution threats, thereby contributing to a lower racial disparity in SARS-CoV-2 infection rates (Mitchell & Popham, 2008; Wolch et al., 2014).

Second, less racial disparity in landscape exposure may lead to less racial disparity in engagement of outdoor physical activities, thereby contributing to a lower racial disparity in SARS-CoV-2 infection rates (Lu, Chen, et al., 2021). A large number of studies have reported that landscape exposure encourages physical activity (Cox et al., 2017; Lu, Zhao, et al., 2021). More frequent physical activity can reduce the risk of obesity (Knobel et al., 2021; Kuo, 2013), cardiovascular diseases (Lanki et al., 2017), stroke (Wilker et al., 2014), systematic inflammation (Bikomeye et al., 2022), and promote immune functioning (da Silveira et al., 2021), making them less vulnerable to SARS-CoV-2 infection (Jiang et al., 2022).

Third, less racial disparity in landscape exposure may lead to less racial disparity in mental stress, thereby contributing to a lower racial disparity in SARS-CoV-2 infection rates. Many studies have shown that landscape exposure significantly reduces mental stress (Jiang et al., 2016; Sullivan & Li, 2021), and stress reduction boosts immune functioning (Kuo, 2015; Segerstrom & Miller, 2004). Studies report that landscape exposure boosts immune functioning by increasing the number of Natural Killer (NK) cells, lymphocytes, and enhanced human NK activities (Li et al., 2010; Li et al., 2008).

Fourth, less racial disparity in landscape exposure may lead to less racial disparity in outdoor social activities, thereby contributing to a lower racial disparity in SARS-CoV-2 infection rates. People with access to nearby landscapes can more easily engage in social activities while remaining at a safe social distance. Compared to indoor settings, outdoor landscape settings allow people to socialize with others at a much lower risk of contracting SARS-CoV-2. Outdoor settings provide high levels of air circulation and more room for social distancing (Leclerc et al., 2020; Miao et al., 2021; Vos et al., 2022).

#### 4.1.2 Why did racial disparity in forest exposure yield the strongest beneficial association with racial disparity in infection rates?

Among all the landscape settings we analyzed, we found that lower racial disparity in population-level exposure to forests was most closely associated with a lower racial disparity in SARS-CoV-2 infection rates. There are two possible explanations. First, forests (including trees along neighborhood streets) are more prevalent than other types of green spaces and distributed more evenly across urban and rural areas in the US (Yang et al., 2018). The relatively even distribution of forests among different neighborhoods provides both black and white population groups more opportunities for nature exposure.

Second, exposure to forests are more effective at reducing mental stress and negative emotions than exposure to other types of landscape (Beil & Hanes, 2013; Jiang et al., 2016), which may make them more effective at improving immune functioning and reducing systematic inflammation, further contributing to a stronger resistance to infection (Kuo, 2015; Li et al., 2010). Third, forests can more effectively capture particulate pollutants than grassland and shrubs because they have a more vertical and complex profile of foliage (Beckett et al., 2000). Exposure to forests is significantly associated with lower incidences of acute respiratory symptoms (Nowak et al., 2006; Nowak et al., 2018) and SARS-CoV-2 infection (Jiang et al., 2022; Lovasi et al., 2008). Lastly, compared to lawn, grassland, and open spaces, forests typically have more dense tree canopy, trunks, and narrow paths, which may encourage social distancing (Jiang et al., 2022). Previous studies have noted that the beneficial impacts of open spaces on SARS-CoV-2 infection (e.g., reduced air pollution and mental stress, enhanced physical activities) may be significantly offset by their detrimental impacts (e.g., increased social contact if people congregate in the open space). This impact may be more profound in dense urban areas (Yang et al., 2022).

#### 4.1.3. Compared to forest inside parks, why does the racial disparity in forests outside parks have a stronger association with racial disparity in infection rates?

There are several possible reasons. Previous studies argue that green spaces mediate health disparities among black and white people due to differences in how these groups interact with green spaces (Iyer et al., 2020). Compared to white people, black people in the US may use parks differently because they have historically been excluded from parks, especially moderate or large parks in suburban and rural areas (Byrne & Wolch, 2009; Wolch et al., 2014). Recent surveys of park users in the US report that black people often cite greater difficulties in using parks than white users, including feeling unwelcome, lacking free time, lacking a private vehicle, and financial stress caused by long-distance travel (Iyer et al., 2020; Wolch et al., 2014). Although black people may have a greater access than white people to urban parks in downtown areas, they experience more obstacles to accessing moderate and large parks in suburban and rural areas. Due to these obstacles, forests inside park may have a weaker impact on infection rates than forest outside park (Larson et al., 2021). In contrast, forests outside parks provide similar access to both black and white people, which allows the positive effect of forests on the SARS-CoV-2 infection can be equally applied to both black and white people.

Moreover, the area of forest outside parks for both races is much larger than the area of forest inside parks (Fig. 3: c and d). This may allow more people to maintain a safe social distance, which may lead to reduced infection (Jiang et al., 2022).

#### 4.1.4 Why is lower racial disparity in low and medium intensity urban areas associated with a lower racial disparity in infection rates?

We found that a higher racial disparity in low or medium development intensity areas were significantly associated with a lower racial disparity in infection rates while high development intensity areas did not yield a significant association with the racial disparity in infection rates. There are several possible explanations for this finding.

First, many studies note that people living in high urban intensity areas have a higher risk of contracting viruses (Acuto et al., 2020; Teller, 2021). People living in these areas are often in close contact with people who may be infected, leading to a greater risk of exposure (Jamshidi et al., 2020; Kokubun & Yamakawa, 2021). Second, studies suggest that people in low and medium urban intensity areas are more likely to engage in outdoor activity than people in high intensity areas, thereby enhancing their immune functioning (Wang et al., 2021). Third, people living in high urban intensity areas have higher levels of stress (Luo & Jiang, 2022). People suffering from stress often have higher levels of cortisol, which can inhibit the production of lymphocytes, which are a type of white blood cell that can help people resist viruses, including SARS-CoV-2 (Jiang et al., 2014; Maydych et al., 2017; Ryden et al., 2009). Lastly, people living in high urban intensity areas may have lower levels of social capital (e.g., Eriksson & Rataj, 2019; Rupasingha et al., 2006). Social capital has been proven to negatively correlate with SARS-CoV-2 infection rates and population density (Kokubun & Yamakawa, 2021). People living in low or moderate density areas may have more equitable access to public spaces and facilities where they can socialize and collaborate. Social capital is critical for actions that require residents to work together to reduce the severity of pandemic, such as wearing masks and social distancing (Watanabe et al., 2022).

### 4.2 Implications

Our findings suggest four major landscape and urban planning implications. First, this study found significant and large racial disparities in both SARS-CoV-2 infection rates and landscape and urban exposures. Furthermore, we found a high level of agreement between the two types of racial disparity. To address this racial disparity, governments should allocate public resources to add vegetation and improve access to natural landscapes in areas with higher concentrations of minority populations. City planners should also work to design less crowded urban environments in these areas to reduce racial disparity in public health outcomes.

Second, we found that different landscape settings and urban development intensities had different associations with SARS-CoV-2 infection rates and with racial disparity in SARS-CoV-2 infection rates. These findings suggest that we should not treat all landscape and urban development types the same when allocating public resources. City administrators, planning and design professionals, and public health professionals need to understand what types of landscape settings and urban development intensities may significantly reduce the risk of SARS-CoV-2 infection.

Third, this study suggests that distance to landscape settings and urban areas impacts their association with SARS-CoV-2 infection. The beneficial effects of forest outside park, pasture/hay, and urban areas with low and moderate development intensity are greatest when they are within a comfortable walking distance (radius buffer under 400m). The effects diminish as distance increases, eventually becoming nonsignificant. These findings suggest that governments should allocate more resources to preserve and develop natural landscapes and areas with low and moderate development intensity within walking distance.

Fourth, this study calls for interdisciplinary cooperation between urban planners and landscape architects. Previous studies typically examine either landscape settings or urban areas because these two types of environments might be essentially complementary, so it is not necessary to examine their effects in the same study, but we found, after controlling for multicollinearity, that both landscape exposure and urban exposure yielded significant associations. Planners should consider landscape and urban exposure as two equally important tools to alleviate risk of SARS-CoV-2 infection and other airborne infectious diseases.

### 4.3 Limitations and future research

This study has two major limitations, which suggest opportunities for future study. First, because we used aggregated SARS-CoV-2 infection data, this study was essentially vulnerable to the ecological fallacy. To largely reduce the risk of ecological fallacy, we used fine-scaled racial map data, population-weighted analysis for landscape and urban exposure at a fine scale, and a within-county comparison of racial disparities in environmental exposures and infection rates. Nevertheless, the environmental exposures and infection rates data at the individual level can improve the accuracy of the findings and should be encouraged to be used in future studies when those data are available.

Moreover, this study identifies a correlational relationship between the racial disparity of landscape and urban exposure and the racial disparity of infection rates. Theoretical and empirical findings from previous studies suggest that there may be a causal relationship. We suggest that future researchers should consider conducting natural experimental studies, environmental intervention studies, or cohort studies to explore more of the possible causal relationship (Jiang et al., 2021).

## Conclusion

The philosopher Georg Hegel said, “The only thing that we learn from history is that we learn nothing from history.” Although many people believe the COVID-19 pandemic is over, we haven’t fully understood it and we haven’t learned enough from it. This nationwide study provides evidence to guide landscape and urban planning to alleviate racial disparities in landscape and urban exposure and SARS-CoV-2 infection rates. Governments, urban planners, landscape planners, and public health professionals should work together to provide more equitable access to certain types of landscape settings and urban areas to reduce risk of infections for all races. In all, we are positive that our findings can contribute to solve similar public health and social crisis in the near or far future.

## Data Availability

All data produced in the present study are available upon reasonable request to the authors

## Notes

### Competing Interest Statement

The authors have declared no competing interest.

### Funding Statement

This work was supported by University Research Committee of The University of Hong Kong (grant: 102010054.088616. 01100.302.01), The University of Hong Kong HKU-100 Scholars Fund, and National Natural Science Foundation of China (No. 51908202).

### Author Declarations

Centers for Disease Control and Prevention (CDC, https://data.cdc.gov/Case-Surveillance/COVID-19-Case-Surveillance-Restricted-Access-Detai/mbd7-r32t)

